# Why we should be sharing our operations: a game theoretic analysis of surgical learning

**DOI:** 10.1101/2025.02.10.25321390

**Authors:** AS Soares, M Chand

## Abstract

**Introduction:** Surgical training has traditionally relied on the master-apprentice model, emphasizing supervised repetition and immediate feedback within the operating room. With the advent of minimally invasive surgical techniques, the capability to record and digitally store surgical procedures has introduced new opportunities for detailed analysis and enhanced feedback mechanisms. Despite this potential, there is a lack of comprehensive systems to analyse recorded surgical procedures at scale.

**Methods:** In this study, we propose a cooperative game-theoretic model to examine the dynamics of surgical training, specifically focusing on the interactions between a master surgeon and an apprentice. The model incorporates both internal knowledge growth—stemming from direct collaboration—and external knowledge growth from supplementary educational resources. A characteristic function is proposed to quantify the utility (knowledge) generated by different coalitions of participants.

**Results:** Our findings demonstrate that collaboration between the master and apprentice leads to a synergistic increase in total knowledge value, surpassing the sum of their individual contributions. The integration of external resources significantly amplifies this effect, showing an exponential impact on knowledge acquisition over time. Proficiency analysis indicates that combining practical experience with structured external learning resources not only accelerates the apprentice’s progression to proficiency but also enhances the overall knowledge within the surgical community.

**Conclusion:** The study underscores the potential of applying game-theoretic principles to optimize surgical education. By quantifying the influence of mentorship quality and external learning resources, we highlight actionable strategies to enhance surgical training outcomes. Embracing a combination of hands-on practice and external resources accelerates individual skill development and enriches the collective knowledge base. We anticipate that this conceptual framework will inform future educational models and encourage the adoption of collaborative and technology-enhanced learning practices in surgery.

## 1 Introduction

Surgical training and practice fundamentally rely on the development of practical skills through the supervised repetition of specific gestures and techniques. Traditionally, this hands-on training model involves a mentor or experienced surgeon providing immediate feedback on the trainee’s performance during operative procedures. The mentor not only assesses the progress of the operation but also assumes control in instances where the trainee demonstrates inadequate progress or encounters complications beyond their current skill level. This apprenticeship-based method has been extensively tested and has served as the cornerstone of surgical education since the advent of modern surgery [1]. One of the most notable changes in recent decades has been the widespread adoption of minimally invasive surgical (MIS) techniques, such as laparoscopic and robotic surgeries. The primary interface for surgeons performing MIS is through imaging systems. This development allows every minimally invasive procedure to be recorded and stored digitally. This capability presents a valuable opportunity for detailed analysis and feedback, which could significantly enhance the training process and improve surgical outcomes. However, despite the potential benefits, there are currently no comprehensive systems in place to analyze each recorded procedure in detail. The manual analysis of surgical videos is exceedingly time-consuming and resource-intensive, representing a substantial missed opportunity for leveraging data-driven insights to advance surgical education and practice [2]. To address this gap, innovative approaches are needed to systematically analyze surgical procedures and provide meaningful feedback to trainees.

Game theory is a mathematical framework for analyzing strategic interactions among rational decision-makers. This field, pioneered by John von Neumann and Oskar Morgenstern in the mid-20th century [3], offers well-defined methodologies to formalize interactions (or “games”) and predict expected outcomes based on the strategies employed by the participants. This field has yielded significant advancements in various domains, including economics, political science, evolutionary biology, and artificial intelligence [4].

In this paper, we propose to apply game theory to the study of surgical practice with the objective of gaining a deeper understanding of the dynamics involved in surgical training and the broader surgical community. By modelling the interactions between mentors and trainees, as well as between different surgical teams, game theory can provide insights into the optimal strategies for skill acquisition, knowledge transfer, and collaborative problem-solving in the operating room. Furthermore, leveraging recorded surgical procedures, we aim to develop analytical tools that can evaluate surgical techniques and decision-making processes through the lens of game-theoretic principles.

## 2 Methods

### 2.1 Game-Theoretic Framework

We developed a cooperative game-theoretic model to analyze knowledge generation in surgical training. This model involves two participants:

- **Master Surgeon (***M*): An experienced surgeon with baseline knowledge.
- **Apprentice (***A*): A less experienced surgeon learning under the master’s guidance.

These participants can form binding commitments and share utilities based on the knowledge they acquire. The **state space** (*τ*) consists of unique surgical cases that influence potential knowledge generation over time.

### 2.2 Directed Acyclic Graph (DAG) Representation

To further elucidate the dynamics between the master surgeon and the apprentice, we incorporated a Directed Acyclic Graph (DAG) into our framework. The DAG visually represents the flow of knowledge and the dependencies between different components of the training process.

**Fig 1.**
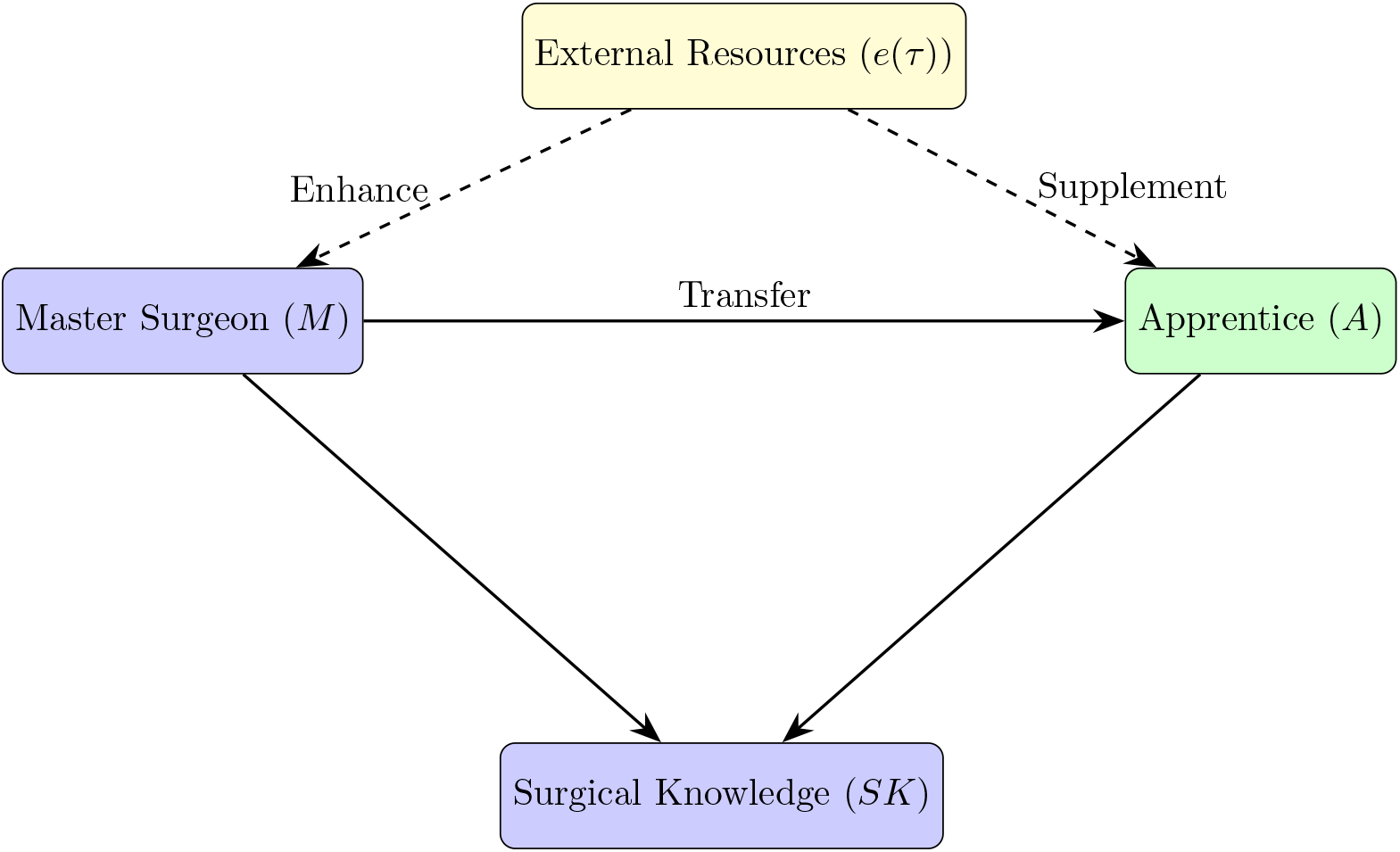
Directed Acyclic Graph (DAG) Representing the Master-Apprentice Relationship

#### Components of the directed acyclic graph

- Master Surgeon (*M*): The central node representing the experienced surgeon.
- Apprentice (*A*): Positioned below the master, indicating the learning relationship.
- External Resources (*e*(*τ*)): External factors that enhance or supplement the knowledge of both the master and the apprentice.
- Surgical Knowledge (*SK*): The aggregate knowledge generated within the surgical community, influenced by both the master and apprentice contributions.
- Arrows:
  - Knowledge Transfer: Indicates the flow of knowledge from the master to the apprentice.
  - Enhancement and Supplement: Show how external resources impact both participants.
  - Contributions to Surgical Knowledge (*SK*): Represent how both the master and apprentice contribute to the overall knowledge pool.

### 2.3 Characteristic Function

The characteristic function, denoted as *v*, quantifies the value or utility that different coalitions (groups) of players can achieve. For our model, the characteristic function is defined as follows:

1. **Empty Coalition (∅):**

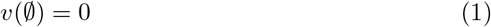
2. **Master Surgeon Alone ({*M*}):**

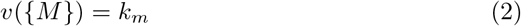
3. **Apprentice Alone ({*A*}):**

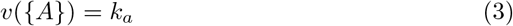
4. **Coalition ({*M, A*}):**

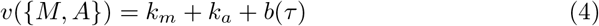

From these equations it follows that: no knowledge is generated when there are no participants, the master surgeon has a baseline knowledge level of *k*_*m*_ and the apprentice has a baseline knowledge level of *k*_*a*_. When both the master and apprentice collaborate, their combined knowledge increases by *b*(*τ*), which represents the additional knowledge generated through cooperation on a specific surgical case *τ* .

### 2.4 Incorporating External and Internal Knowledge Growth

To accurately model the evolving knowledge within a master-apprentice coalition, we incorporate both internal collaborative knowledge growth and external educational resources over time. The characteristic function, representing the coalition’s knowledge value, is modified to reflect this:

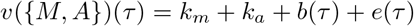

where:

- *k*_*m*_ and *k*_*a*_ represent the baseline knowledge levels of the master surgeon and apprentice, respectively.
- *b*(*τ*) represents the growth of knowledge from repeated collaboration on cases, progressing at a linear rate.
- *e*(*τ*) represents knowledge growth influenced by external resources, modelled with an exponential growth rate.

#### 2.4.1 Internal Knowledge Growth

The internal knowledge growth function, *b*(*τ*), quantifies the incremental knowledge gained through the master-apprentice collaboration alone. Modeled as a linear function, this term captures the cumulative experience over time:

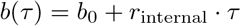

where:

- *b*_0_ is the initial knowledge gain from collaboration at *τ* = 0,
- *r*_internal_ is the linear growth rate of knowledge from ongoing collaboration,
- *τ* represents the time step or number of cases completed together.

#### 2.4.2 External Knowledge Growth

To account for the influence of external educational resources such as third-party training programs, virtual platforms, and expert consultations, we define an external knowledge function *e*(*τ*). This function grows exponentially to capture the increasing impact of external resources over time:

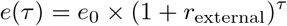

where:

- *e*_0_ is the initial level of external knowledge at *τ* = 0,
- *r*_external_ is the exponential growth rate representing the impact of accumulating external knowledge sources.

This function allows parameterized control over the growth rates for internal and external knowledge, facilitating analysis of how collaboration and external resources impact knowledge over time. By adjusting these parameters, various scenarios of coalition knowledge development can be visualized and analyzed effectively.

### 2.5 Model Assumptions and Limitations

Our model is based on several key assumptions:

- **Perfect Information Sharing:** Both the master and apprentice have complete and accurate information about each other’s knowledge and activities.
- **Rational Behaviour:** All participants act rationally to maximize their utility.
- **Measurable Knowledge Outcomes:** The knowledge generated can be quantitatively measured.
- **Transferable Utility:** The utility (benefit) derived from knowledge can be freely distributed between the master and apprentice.

**Limitations:** These assumptions may restrict the model’s applicability in real-world scenarios where information asymmetry exists or participants do not behave rationally.

### 2.6 Proficiency Analysis

We analyzed proficiency based on the collected data and specific training requirements, focusing on three key metrics:

#### 2.6.1 Overall Proficiency

**Proficiency (Proficiency**(*p*)) is calculated by summing the knowledge units generated across all performed procedures:

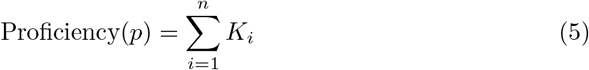

- *K*_*i*_: Knowledge units generated from procedure *i*.
- *n*: Total number of procedures performed.

#### 2.6.2 Time to Proficiency

**Time to Proficiency (***T*_*p*_) estimates the duration required to reach a predefined knowledge threshold:

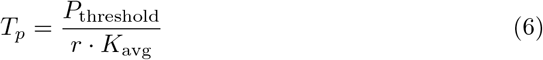

Where:

- *P*_**threshold**_: The required proficiency level measured in knowledge units.
- *r*: Number of procedures performed per unit of time.
- *K*_**avg**_: Average knowledge units generated per procedure.

#### 2.6.3 Community Knowledge Impact

**Total Knowledge within the Surgical Community (***SK*) is the aggregate knowledge generated by all practicing surgeons, considering both internal practice and external resources:

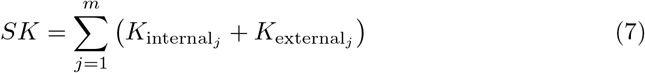

Where:

- *m*: Number of practicing surgeons.
- 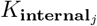: Knowledge generated through the direct practice of surgeon *j*.
- 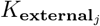: Knowledge acquired by surgeon *j* from external resources.

This metric provides insight into the overall knowledge landscape within the surgical community, highlighting the contributions of individual practice and external learning opportunities.

### 2.7 Model analysis

The model presented before was subject to interrogation by varying the inputs, in order to identify the contributions of the relevant factors using python code, available here. This manuscript was prepared with the assistance of ChatGPT, model 4o and o1 preview, used between October and November 2024.

## 3 Results

### 3.1 Game-Theoretic Model Outcomes

Our cooperative game-theoretic model demonstrates that collaborative knowledge generation between the master surgeon (*M*) and apprentice (*A*) can produce a synergistic increase in total knowledge value. When both participants work together, the characteristic function *v*({*M*,*A*}) = *k*_*m*_ + *k*_*a*_ + *b*(*τ*) shows that the combined knowledge generated exceeds the individual contributions of each surgeon.

The results indicate that this cooperative framework maximizes value through extended collaboration, as the function *b*(*τ*) grows linearly with time and case exposure. Optimal outcomes are achieved when the participants are continuously engaged in varied and challenging surgical cases, supporting rapid knowledge accumulation. This dynamic aligns with real-world mentorship, where repeated exposure and guidance in complex scenarios produce proficiency at a faster rate than isolated practice.

We performed a sensitivity analysis varying three parameters: master surgeon knowledge (*k*_*m*_), apprentice knowledge (*k*_*a*_), and knowledge gained from internal collaboration (*r*_internal_). The analysis showed identical slopes for different levels of master and apprentice knowledge, as illustrated in figures 2 and 3. Higher internal knowledge generation resulted in steeper knowledge accumulation curves, shown in figure 4.

**Fig 2.**
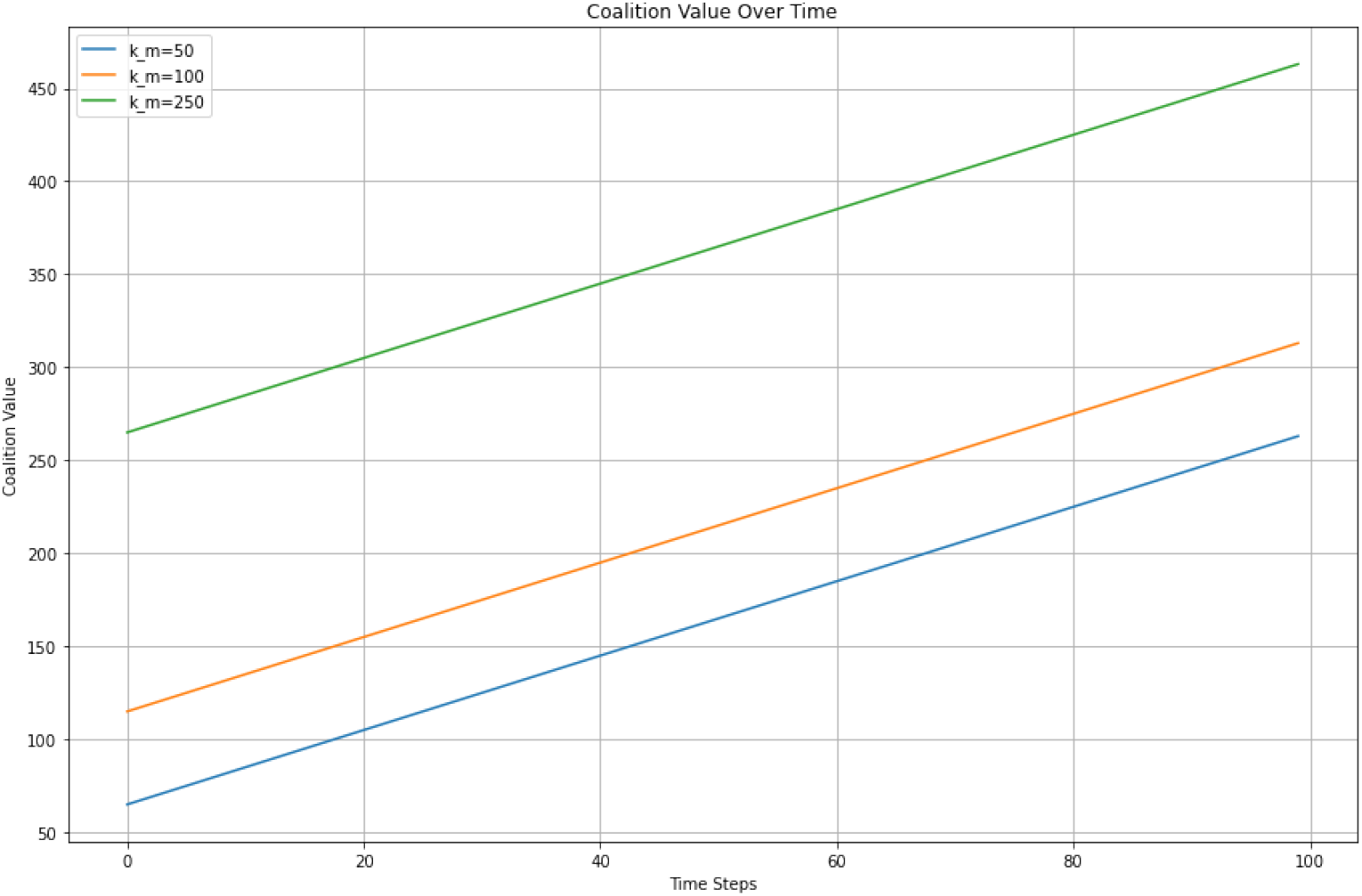
Sensitivity analysis considering different values for master surgeon knowledge *k*_*m*_ (50, 100, 250)

**Fig 3.**
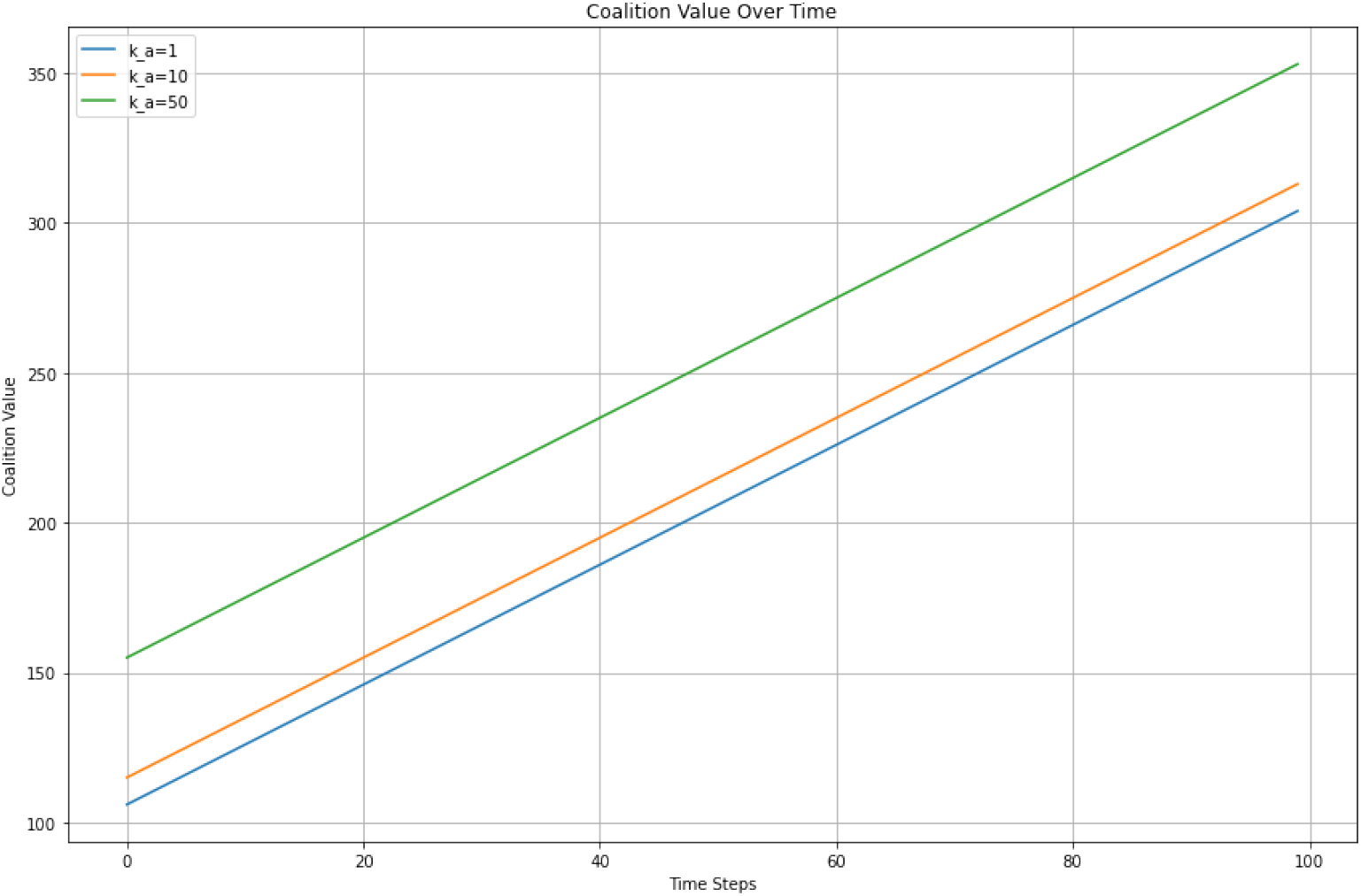
Sensitivity analysis considering different values for apprentice knowledge *k*_*a*_ (1, 10, 50)

**Fig 4.**
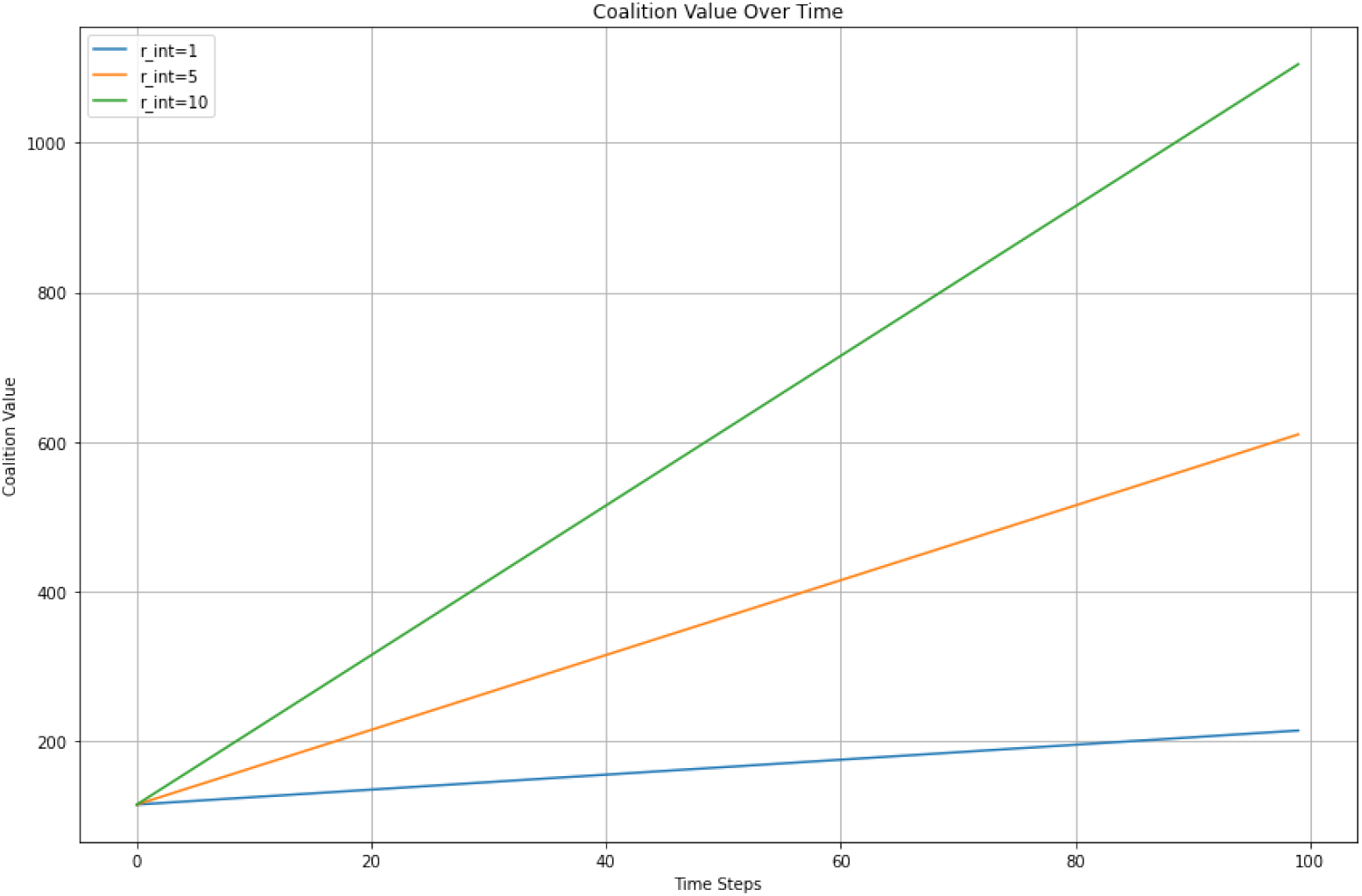
Sensitivity analysis considering different values for knowledge from internal collaboration *r*_internal_ (1, 5, 10)

### 3.2 Incorporating External Knowledge

External resources (*e*(*τ*)), represented in the DAG as external factors enhancing both *M* and *A*, play a significant role in boosting the knowledge growth rate for the coalition. The external knowledge growth function *e*(*τ*) = *e*_0_ × (1 + *r*_external_)^*τ*^ reflects an exponential impact over time, highlighting the compound benefits of supplemental learning tools, online training modules, and external consultations. This exponential growth contributes to *v*({*M*,*A*})(*τ*) = *k*_*m*_+*k*_*a*_ +*b*(*τ*)+*e*(*τ*), significantly enhancing the apprentice’s progression and allowing the master to incorporate advanced, updated practices, as can be seen in fig. 5.

**Fig 5.**
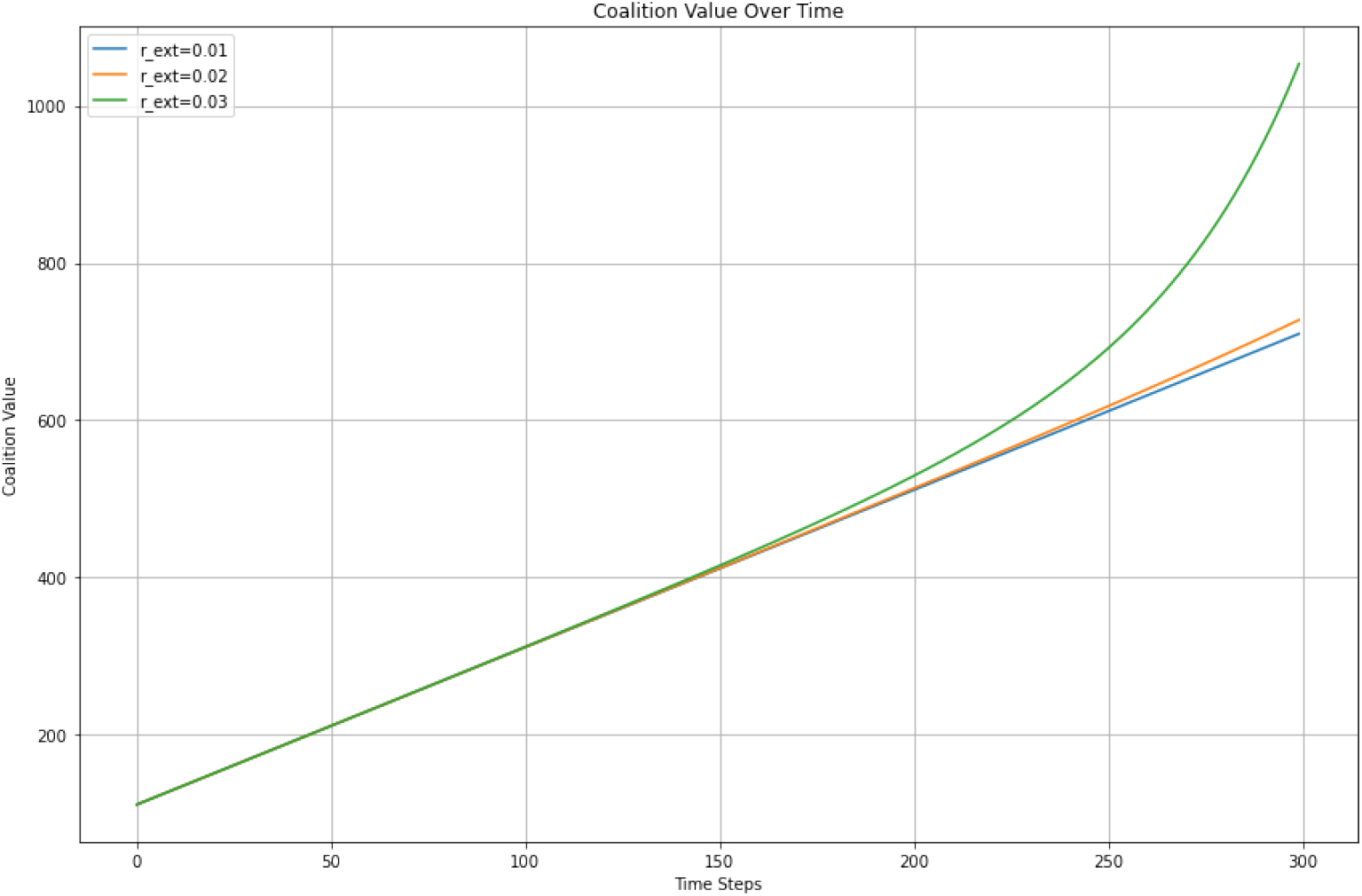
Sensitivity analysis considering different values for external knowledge growth *r*_external_ (0.01, 0.02, 0.03)

The results underscore that the value derived from external resources is maximized when these resources complement hands-on practice. When both participants use these resources systematically, they achieve higher knowledge levels more efficiently, reducing the overall time to reach critical proficiency milestones. This setup suggests that a structured integration of external learning provides an optimal strategy for training within surgical education, as it builds a foundation of continuously reinforced skills.

### 3.3 Proficiency Analysis

#### 3.3.1 Overall Proficiency

Calculating total proficiency as the sum of knowledge units generated from all procedures, Proficiency 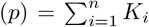, the results show that assuming a constant level of knowledge generation, eventually any level of proficiency will be reached. As the apprentice (*A*) engages in cases with the guidance of the master (*M*), the cumulative knowledge *p* approaches higher levels over time, emphasizing the incremental effect of each procedure.

#### 3.3.2 Time to Proficiency

The model calculates the time required to reach a pre-defined knowledge threshold, 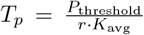. Results show that frequent case exposure and increased knowledge units per procedure accelerate proficiency. As can be seen in fig. 6, for any given level of proficiency, higher *r*_external_ values will decrease the time to proficiency. Pairing hands-on practice with strategic external learning resources reduces *T*_*p*_, supporting the hypothesis that a combination of internal practice and external resources optimizes the time to achieve surgical competency.

**Fig 6.**
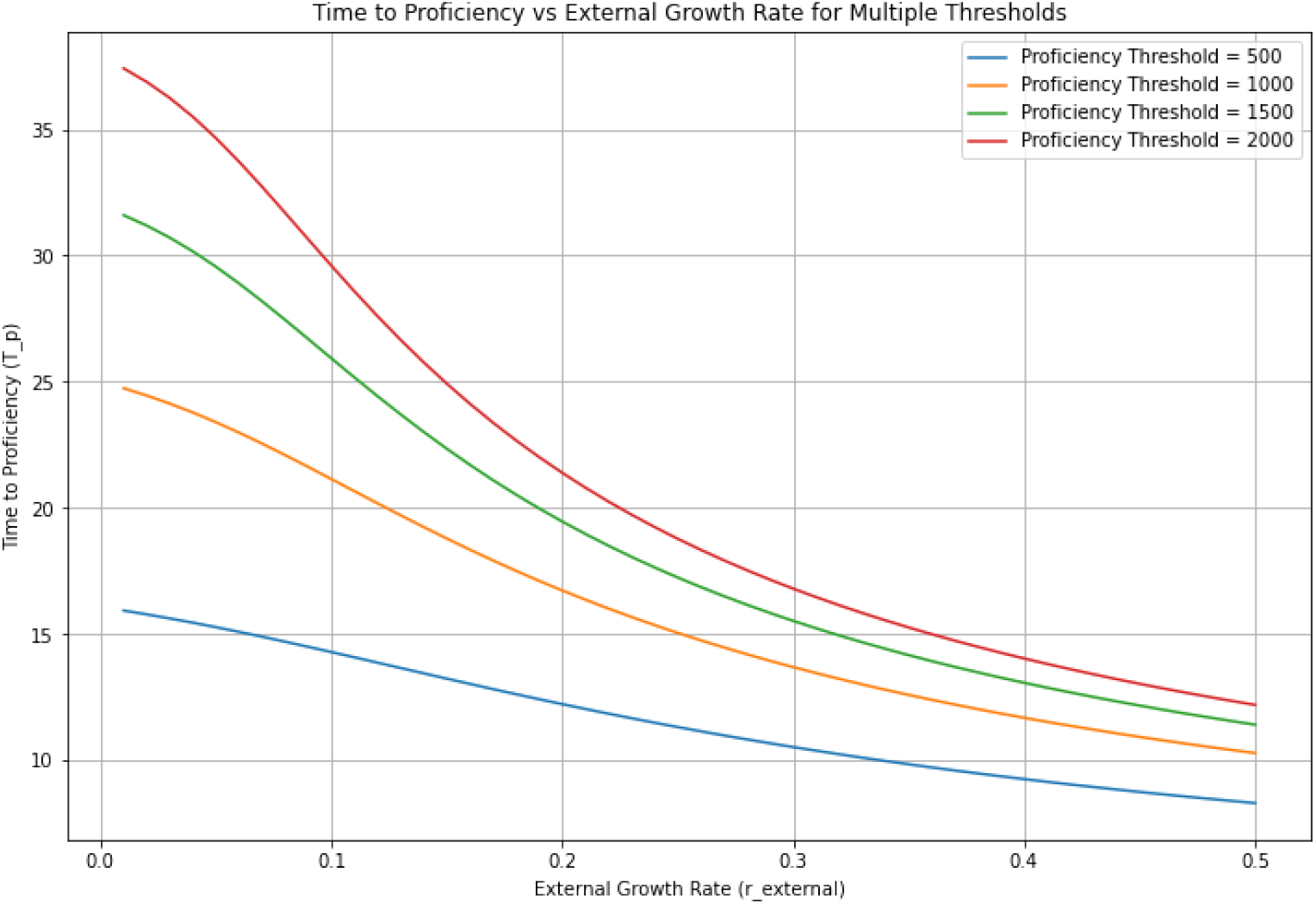
Time to proficiency according to *r*_external_. For any given proficiency level, higher values of *r*_external_ will lead to reaching that level in a smaller number of time steps

#### 3.3.3 Community Knowledge Impact

By evaluating the collective knowledge within the surgical community, 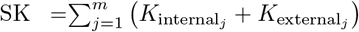, we can measure how individual surgeons contribute to the broader knowledge landscape. The analysis reveals that the integration of external resources, combined with each surgeon’s personal practice, enhances the total community knowledge as can be seen in fig. 7. This collective growth underscores the broader implications of incorporating structured training pathways and external learning tools across the community, where individual advancements drive overall improvement in surgical practices.

**Fig 7.**
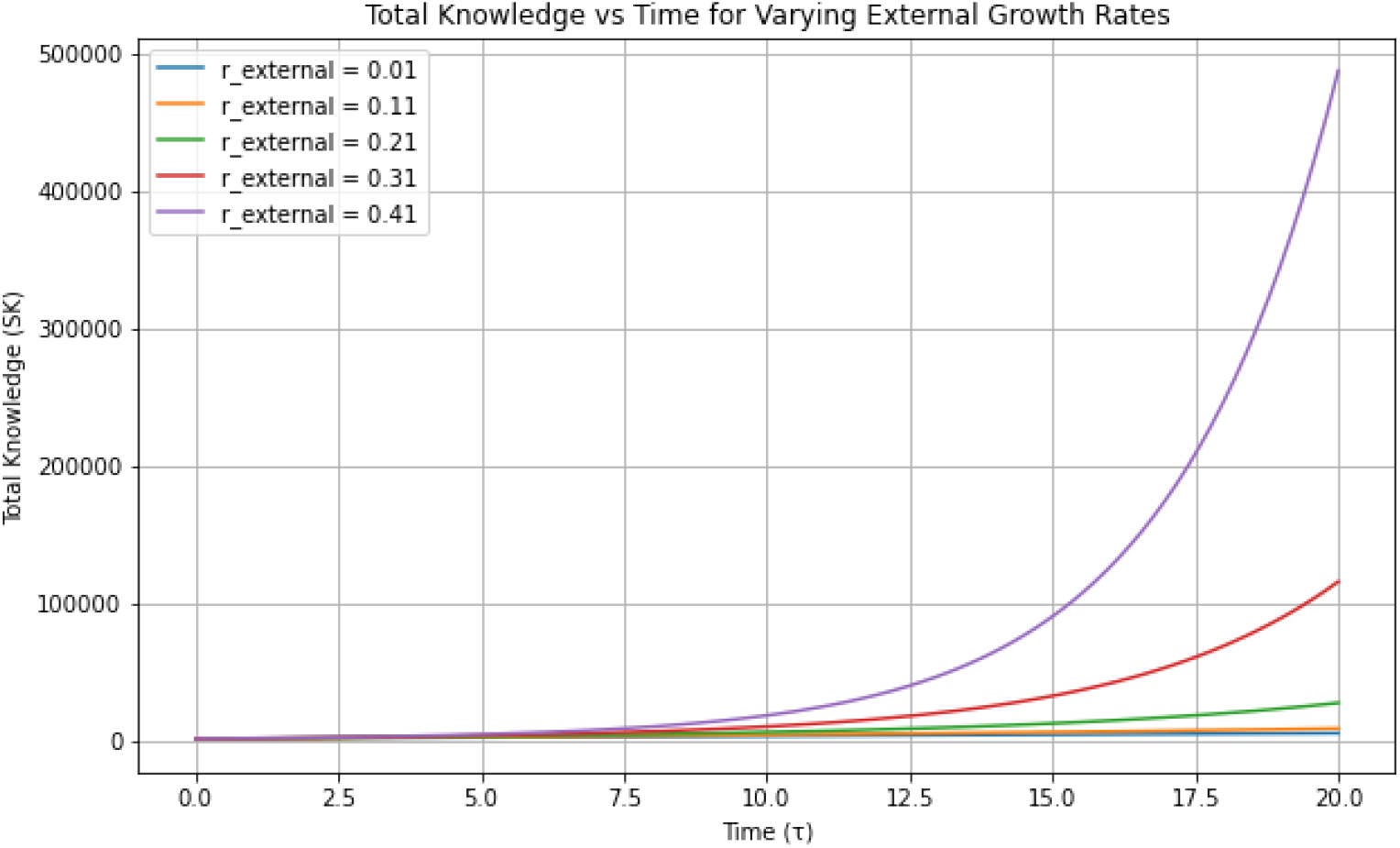
Total knowledge over time for different *r*_external_ values.

In summary, the results validate that a cooperative game-theoretic approach maximizes knowledge acquisition through collaborative engagement. External resources amplify this effect, and proficiency metrics confirm that sustained collaboration and supplemental learning expedite surgical skill development, thus benefiting both individual practitioners and the surgical community as a whole.

## 4 Discussion

This study introduces a cooperative game-theoretic model to analyze the dynamics of surgical learning between a master surgeon (*M*) and an apprentice (*A*). By incorporating key parameters—the knowledge levels of the master (*k*_*m*_) and apprentice (*k*_*a*_) surgeons, the gain in knowledge from internal collaboration (*r*_internal_), and the gain from external elements (*r*_external_) — we demonstrate how these factors can influence knowledge acquisition over time. Our findings indicate that apprentices with higher internal learning rates accumulate more knowledge within the same timeframe, emphasizing the importance of effective mentorship and hands-on experience. Furthermore, the integration of external knowledge resources yields a disproportionate increase in knowledge acquisition, substantially reducing the time required to achieve proficiency (*T*_*p*_). Over time, this leads to a significant expansion of the total surgical knowledge pool (*SK*), benefiting not only individual surgeons but also the broader surgical community.

The theoretical framework presented allows for a nuanced interrogation of surgical training components that are well-recognized by practitioners yet challenging to quantify. It highlights the impact of varying levels of master and apprentice knowledge and underscores the critical role of learning rates in skill acquisition. Notably, the model illustrates that significant gains can be achieved by incorporating external knowledge, which can be aggregated across diverse settings.

Considering that an estimated 313 million surgical procedures are performed globally each year [5], and approximately 15.8% of these are minimally invasive surgeries[6], there exists a vast potential for enhancing surgical practice through shared learning. This translates to 49 450 000 minimally invasive procedures annually that could contribute to a collective learning repository. The exponential impact of external knowledge, as demonstrated by our model, suggests that embracing tools such as surgical video libraries, online modules, and virtual simulations could revolutionize surgical education. By systematically integrating these resources, surgeons can accelerate proficiency development, reduce the time to achieve competency, and contribute to the cumulative advancement of surgical techniques.

The model aligns with contemporary educational paradigms that emphasize blended learning and continuous professional development. It supports the notion that combining practical experience with structured external learning optimizes training outcomes. This approach is particularly relevant in the context of modern surgical practice, where technological advancements and procedural innovations necessitate ongoing learning.

### 4.1 Limitations and Future Directions

While our model offers valuable insights, it is grounded in several simplifying assumptions that warrant consideration. We assume that knowledge generation is a quantifiable output from the interaction between a master surgeon and an apprentice, which, in reality, is difficult to measure precisely. The model abstracts complex educational and interpersonal dynamics into quantifiable parameters, potentially oversimplifying the multifaceted nature of surgical learning.

Moreover, the tools required to effectively incorporate external knowledge into surgical practice are still evolving. There is a notable lack of large, accessible datasets with standardized outcomes that can be utilized by surgeons globally. The majority of surgical procedures are not routinely recorded; estimates suggest that approximately 20% of surgeons do so in the United Kingdom[7]. This figure is likely to be much lower in other countries. Additionally, there is a significant gap in the ability to analyse surgical videos at scale. Current computational methods for video analysis are not yet sophisticated enough to provide actionable feedback without substantial manual input [2].

Addressing these limitations requires concerted efforts to promote the recording and sharing of surgical procedures while ensuring patient confidentiality and data security. Developing standardized protocols for video capture and sharing, alongside advancements in artificial intelligence and machine learning for automated video analysis, could bridge the existing gaps. Future research should focus on validating the model with empirical data, exploring the impact of different types of external resources, and refining the parameters to better reflect the complexities of surgical education.

## 5 Conclusion

The cooperative game-theoretic model details the synergistic effects of collaboration and external knowledge integration in surgical training. By quantifying the influence of mentorship quality and external learning resources on knowledge acquisition and proficiency attainment, actionable strategies to optimize surgical education are highlighted. Embracing a combination of hands-on practice and external resources not only accelerates individual surgeon development but also enriches the collective knowledge base of the surgical community. We anticipate that this conceptual framework will inform future educational models and encourage the adoption of collaborative and technology-enhanced learning practices in surgery.

## Data Availability

All data produced are available online at a dedicated github repository.

https://github.com/asampaiosoares/game_theory_surgery

